# A Multi-center Study of COVID-19 with Multivariate Prognostic Analysis

**DOI:** 10.1101/2020.09.26.20202234

**Authors:** Wen Zeng, Xin Feng, Jie Huang, Chuan Du, Dongming Qu, Xiang Zhang, Jianquan Zhang

## Abstract

**Purpose:** Coronavirus disease (COVID-19) pandemic is now a global health concern. However, there is no detailed analysis of the factors related to patients’ improvement.

**Patients and methods:** We compared the clinical characteristics, laboratory findings, CT images, and treatment of COVID-19 patients from two different cities in China. One hundred and sixty-nine patients were recruited from January 27 to March 17, 2020 at five hospitals in Hubei and Guangxi. They were divided into four groups according to age and into two groups according to presence of comorbidities. Multivariate statistical analyses were performed for the prognosis of the disease.

**Results:** Fifty-two patients (30.8%) had comorbidities, and the percentage of critical COVID-19was higher in the comorbidities group (11.6%vs.0.9%, p<0.05). Older patients had higher proportion of severe or critical disease. The results showed that lymphocyte count was significantly associated with the number of days from positive COVID-19 nucleic acid test to negative test; number of days from onset of symptoms to confirmation of diagnosis was significantly associated with the time it took for symptoms to improve; and number of days from onset of symptoms to confirmation of diagnosis and disease severity were significantly associated with chest computed tomography improvement.

**Conclusions:** Age, comorbidities, lymphocyte count, and SpO_2_ may predict the risk of severity of COVID-19. Early isolation, early diagnosis, and early initiation of management can slow down the progression and spread of COVID-19.

**Key Points:** Age and comorbidities can predict the risk of severity of COVID-19, Lymphocyte count and SpO_2_ may predict the risk of severity of COVID-19. Early isolation, Early diagnosis can slow down the progression of COVID-19

## 1. Introduction

Coronavirus disease 2019 (COVID-19), which is caused by severe acute respiratory syndrome coronavirus2 (SARS-CoV-2), has spread throughout the world, posing a critical threat to global health. COVID-19 has been declared as a public health emergency by the World Health Organization, and 3,175,207 laboratory-confirmed infections had been reported globally by May 1, 2020.1 Several studies have described the clinical characteristics and epidemiology of COVID-19.2-6 The novel corona virus has caused clusters of severe respiratory illness, similar to severe acute respiratory syndrome coronavirus.2,5 Previous studies have also summarized the early clinical, laboratory, and computed tomography (CT)findings of COVID-19 pneumonia.3,6 A multi-center study of clinical features showed that multiple organ dysfunction and impaired immune function were the typical characteristics of severe and critical disease, and advanced age (≥75 years) was a risk factor for mortality.4 However, there is no detailed analysis of the factors related to patients’ improvement. In this study, we analyzed the clinical features, laboratory findings, CT images, outcomes, and therapies (including antiviral drugs, antibacterial drugs, and corticosteroids)in 169 COVID-19 cases from three cities (Nanning and Guilin in Guangxi Province, and Wuhan in Hubei Province).We also analyzed the factors related to patients’ improvement.

## 2. Methods

### 2.1. Study design and participants

Patients who were ≥18 years old were recruited for this multi-center retrospective study from five hospitals designated for the treatment of COVID-19 in China, namely Guangxi Medical University First Affiliated Hospital and The Fourth People’s Hospital of Nanning City, both in Nanning; Nanxishan Hospital of Guangxi Zhuang Autonomous Region, in Guilin, Guangxi Province; and Wuhan Huangpi District Hospital of Traditional Chinese Medicineand Hubei No.3 People’s Hospital of Jianghan University, bothin Wuhan, Hubei Province. The recruitment period was from January 27 to March 17, 2020. All patients enrolled in this study were diagnosed with COVID-19 according to the diagnostic criteria from the Chinese Clinical Guidance for COVID-19 Pneumonia Diagnosis and Treatment(7th edition), which was produced by the National Health Commission of China.7 The study was approved by the Research Ethics Commission of Guangxi Medical University First Affiliated Hospital approval number: 2020(KY-E-06).

We grouped the patients according to their ages into group A (18–44 years), group B (45–64 years), group C (64–74 years), and group D (>75 years).Analysis of prognosis of improvement included three aspects: number of days from positive COVID-19 nucleic acid test to negative test, number of days from onset of illness(the first day of presenting with COVID-19-related symptoms, such as fever, cough, and diarrhea)to improvement in symptoms, and improvement on the second CT scan. The primary outcomes were improvement and death.

### 2.2. Definitions

According to the Chinese Clinical Guidance for COVID-19 Pneumonia Diagnosis and Treatment (7th edition), COVID-19 is classified by severity into mild, moderate, severe, and critical types.7

### 2.3. Data collection

The medical records of the recruited COVID-19 patients were reviewed and general information, epidemiological data, and data on clinical presentation, laboratory tests, chest CT scan findings, treatment modalities, and outcome were collected. Clinical data collection was completed on March 17, 2020. Du Chuang and Huang Jie cross-checked the data. The local Center for Disease Control (CDC) laboratories made definitive diagnosis of COVID-19 by analyzing throat-swab specimens from the upper respiratory tract or feces. Real-time reverse transcription polymerase chain reaction (RT-PCR) was used to confirm COVID-192 and exclude other viral infections. All patients underwent chest CT scan. Two attending radiologists were invited to interpret all chest CT scans independently, and they were blinded to the clinical information of each patient. The following CT features were assessed: distribution of lesions (peripheral, along the bronchi, random, or diffuse), number of lobes involved (one, two or three, or four or five), shape of lesions (patchy or nodular), appearance (ground-glass opacity [GGO], consolidation, or GGO with consolidation), specific focal signs (vascular thickening, crazy paving pattern, and fibrosis), and extrapulmonary manifestations (mediastinal and hilar lymph node enlargement, pleural effusion, or pleural thickening).In case of discordance between the interpretations of the two radiologists, the opinion of a third deputy chief physician radiologist was sought to reach a final decision. Data on follow-up, prognosis, and treatment were updated on April 24, 2020.

### 2.4. Statistical analyses

Continuous variables were expressed as medians with interquartile ranges (IQR), and categorical variables were reported as frequencies and percentages. Differences between groups were compared using the independent samples t-test or Mann-Whitney test. The single factor analysis of variance (ANOVA) or Kruskal-Wallis H test was used, as appropriate, to compare the difference among the four groups into which patients were divided based on severity of the disease. Least significant difference (LSD) or Dunn’s test was used for post hoc comparisons. Categorical data were analyzed either with Pearson Chi-square test or with Fisher’s exact test. Two-tailed tests were performed to determine statistical significance at the 5% level. Bonferroni correction was used for pairwise comparison.

Univariable and multivariable analyses for factors associated with the time for nucleic acid test to turn negative and the time of improvement of symptoms were performed with multiple linear regression analyses. Univariable and multivariable analyses for factors that are associated with chest CT improvement were done with Kaplan-Meier method and Cox proportional hazards regression model. Univariable and multivariable analyses for risk factors for disease severity and the relationship between age and disease severity were performed with logistic regression analyses. All data analyses were carried out using IBM Statistical Package for the Social Sciences for Windows, version 25 (SPSS Inc., Chicago, Illinois), and graphs were created using GraphPad Prism 7.0 software (GraphPad Software Inc., San Diego, CA, USA). P value less than 0.05 was considered statistically significant.

## 3. Results

The median age of the patients was 52.8 years (IQR, 19–82). Male patients accounted for 46.7% of all patients. Based on severity, 12 patients (7.1%) had mild COVID-19,137(81.1%) patients had moderate type, 13(7.7%) patients had severe type, and 7(4.1%) patients had critical type. The median number of days from the onset of illness to admission was 11.6 days (IQR, 18–92).Fifty-two patients had comorbidities, including hypertension, cardiovascular disease, diabetes, malignancy, cerebrovascular disease, chronic obstructive pulmonary disease(chronic obstructive pulmonary disease),chronic nephropathy, immunosuppression, and others. The most common morbidity was hypertension(17.8%). Clinical symptoms included fever, cough, sputum production, dry cough, pharyngalgia, shortness of breath, hemoptysis, myalgia, digestive symptoms, hypodynamia, and others. Fever was the most common symptom (76.9%), followed by dry cough (68%). Among the 169 patients, the lymphocyte count was decreased in 111 (65.68%) patients (median=1.24 (IQR=0.18–4.11) ×109/L). C-reactive protein (CRP) increased in 102 (65.39%) patients, lactate dehydrogenase (LDH) increased in 52 (33.33%) patients, and D-dimer increased in 55 (35.95%) patients. Alanine transaminase or aspartate aminotransferase was elevated in 56 patients (33.17%). Serum creatinine was elevated in 5 patients (2.96%), 4 patients had chronic nephropathy, and 1 patient’s renal function returned to normal after treatment. One hundred and fifty-four (90.1%) patients received antiviral therapy within the first 3 days. Antivirals used included lopinavir, tonavir, abidor, ribavirin, ganciclovir, and oseltamivir. Most patients (53.9%) received antibacterial therapy, including moxifloxacin, cephalosporins, and azithromycin. Forty-eight (28.4%) patients received corticosteroids. One hundred and sixty-five (97.60%) patients improved, while four patients died.

Among the 169 patients, 121 were from hospitals in Wuhan, while 48 were from hospitals in Guangxi. We compared the clinical data between the two groups. The Guangxi group had fewer days from onset of illness to admission (5.24:14.09) (p<0.05). Cough and sputum production were more common in the Wuhan group(p<0.05). The total protein and albumin in patients from Wuhan group were lower, while LDH was higher (p<0.05). The four patients who died were all in Wuhan group (Table 1).

**Table 1:**
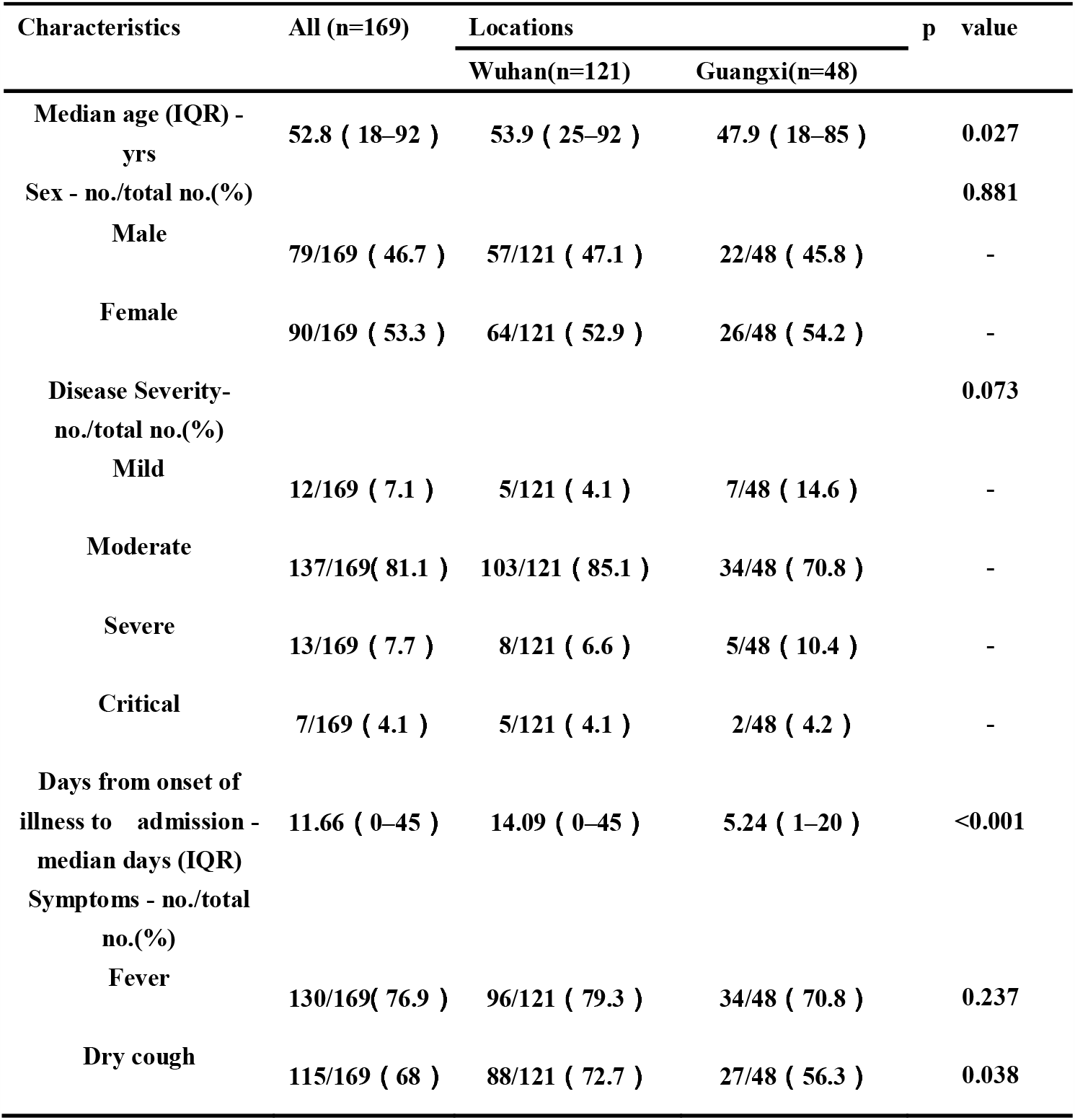
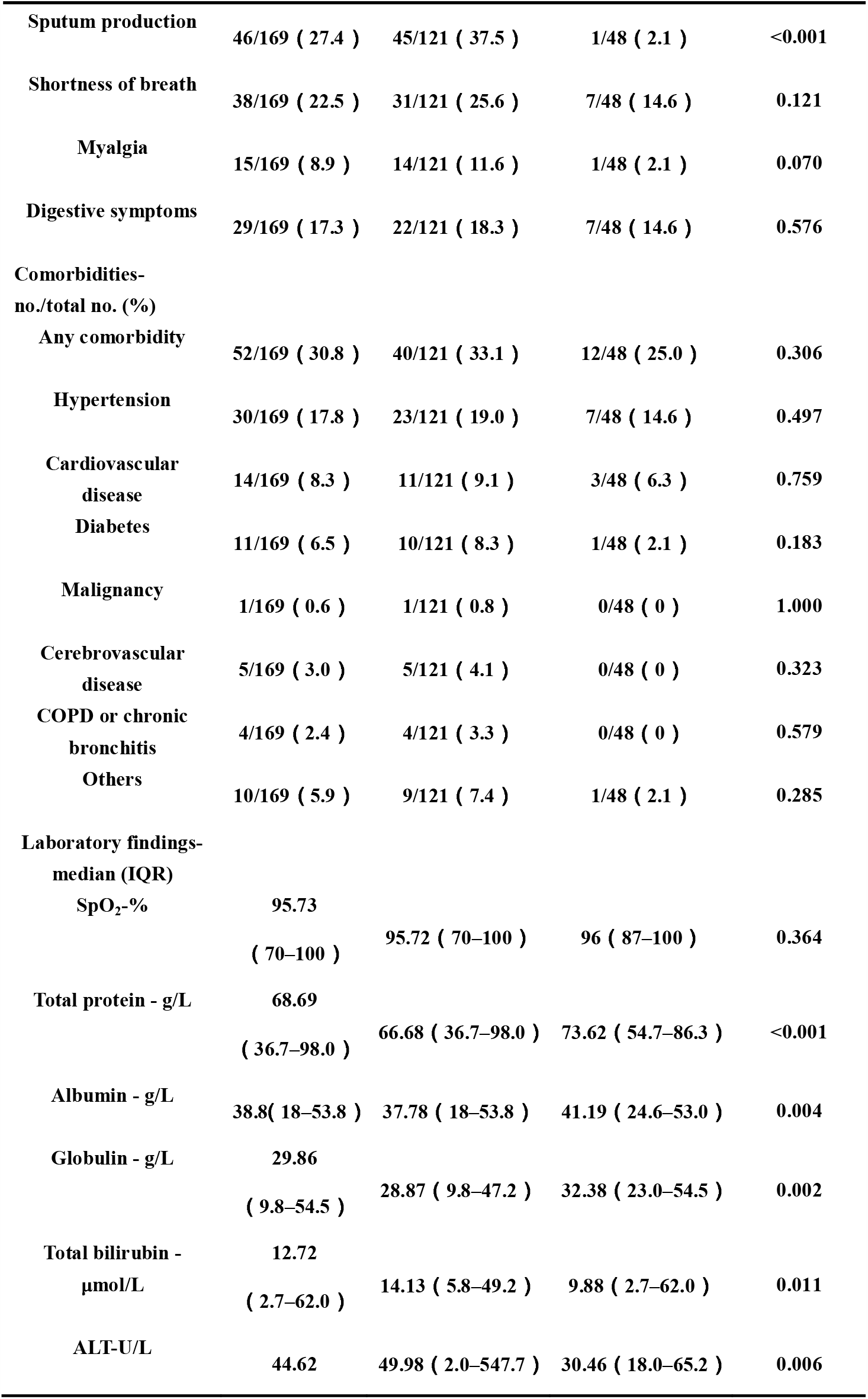
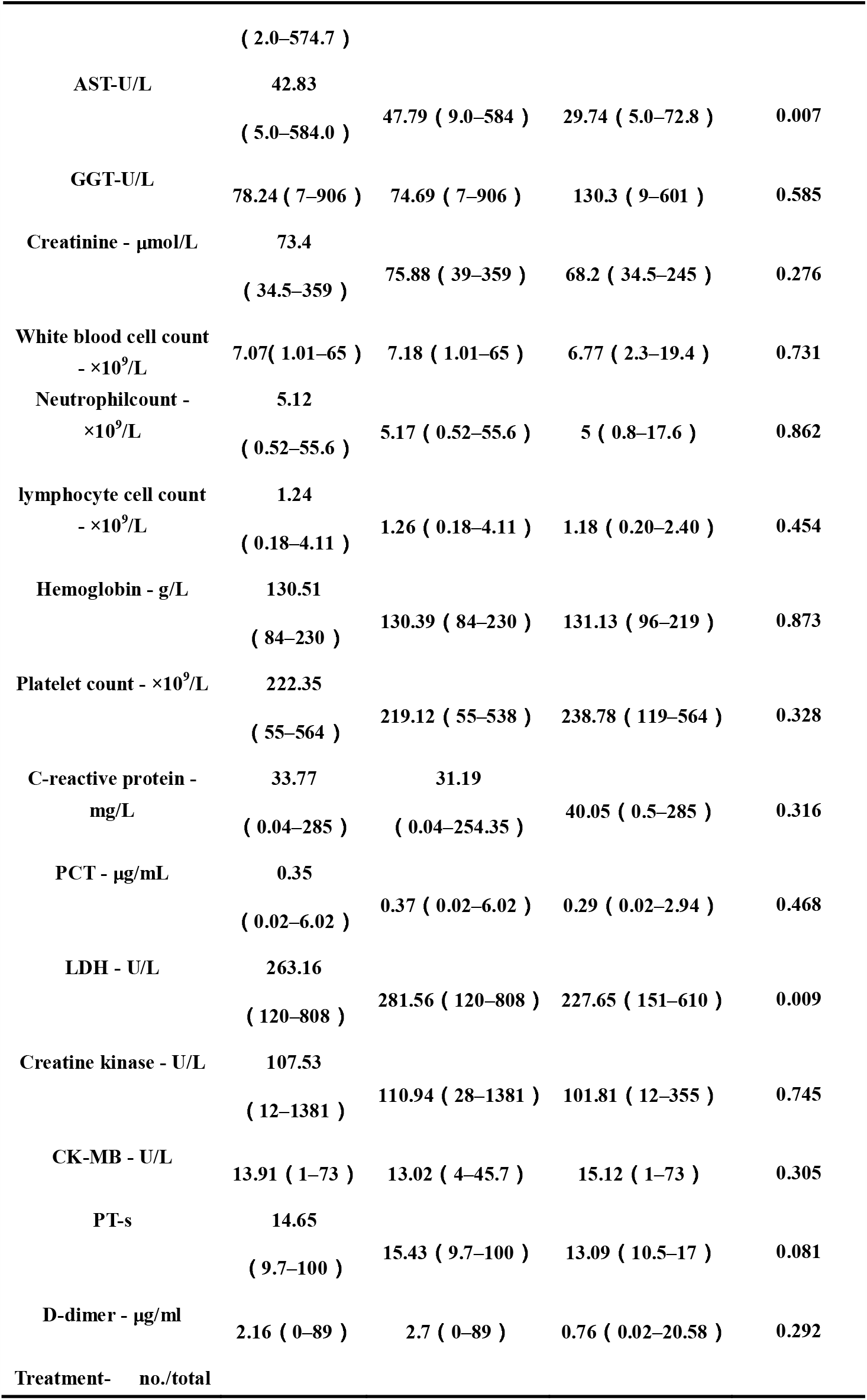
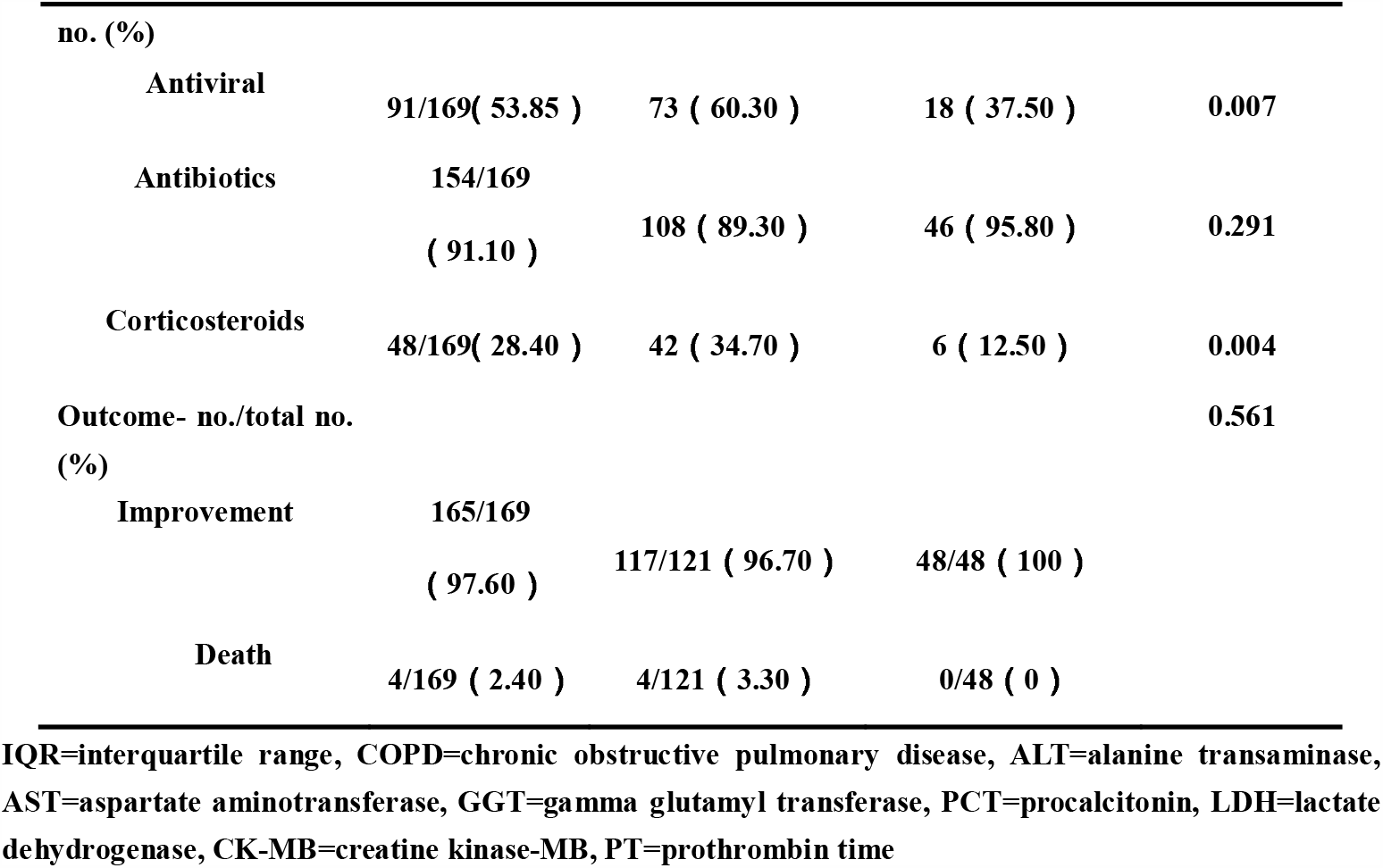
Clinical features of study subjectsin different locations

| Characteristics | All (n=169) | Locations |  | p value |
| --- | --- | --- | --- | --- |
|  |  | Wuhan(n=121) | Guangxi(n=48) |  |
| **Median age (IQR) - yrs** | **52.8 ( 18–92 )** | **53.9 ( 25–92 )** | **47.9 ( 18–85 )** | **0.027** |
| **Sex - no./total no.(%)** |  |  |  | **0.881** |
| **Male** | **79/169 ( 46.7 )** | **57/121 ( 47.1 )** | **22/48 ( 45.8 )** | **-** |
| **Female** | **90/169 ( 53.3 )** | **64/121 ( 52.9 )** | **26/48 ( 54.2 )** | **-** |
| **Disease Severity- no./total no.(%)** |  |  |  | **0.073** |
| **Mild** | **12/169 ( 7.1 )** | **5/121 ( 4.1 )** | **7/48 ( 14.6 )** | **-** |
| **Moderate** | **137/169 ( 81.1 )** | **103/121 ( 85.1 )** | **34/48 ( 70.8 )** | **-** |
| **Severe** | **13/169 ( 7.7 )** | **8/121 ( 6.6 )** | **5/48 ( 10.4 )** | **-** |
| **Critical** | **7/169 ( 4.1 )** | **5/121 ( 4.1 )** | **2/48 ( 4.2 )** | **-** |
| **Days from onset of illness to admission - median days (IQR)** | **11.66 ( 0–45 )** | **14.09 ( 0–45 )** | **5.24 ( 1–20 )** | **<0.001** |
| **Symptoms - no./total no.(%)** |  |  |  |  |
| **Fever** | **130/169 ( 76.9 )** | **96/121 ( 79.3 )** | **34/48 ( 70.8 )** | **0.237** |
| **Dry cough** | **115/169 ( 68 )** | **88/121 ( 72.7 )** | **27/48 ( 56.3 )** | **0.038** |
| **Sputum production** | **46/169 ( 27.4 )** | **45/121 ( 37.5 )** | **1/48 ( 2.1 )** | **<0.001** |
| **Shortness of breath** | **38/169 ( 22.5 )** | **31/121 ( 25.6 )** | **7/48 ( 14.6 )** | **0.121** |
| **Myalgia** | **15/169 ( 8.9 )** | **14/121 ( 11.6 )** | **1/48 ( 2.1 )** | **0.070** |
| **Digestive symptoms** | **29/169 ( 17.3 )** | **22/121 ( 18.3 )** | **7/48 ( 14.6 )** | **0.576** |
| **Comorbidities-**<br>**no./total no. (%)** |  |  |  |  |
| **Any comorbidity** | **52/169 ( 30.8 )** | **40/121 ( 33.1 )** | **12/48 ( 25.0 )** | **0.306** |
| **Hypertension** | **30/169 ( 17.8 )** | **23/121 ( 19.0 )** | **7/48 ( 14.6 )** | **0.497** |
| **Cardiovascular**<br>**disease** | **14/169 ( 8.3 )** | **11/121 ( 9.1 )** | **3/48 ( 6.3 )** | **0.759** |
| **Diabetes** | **11/169 ( 6.5 )** | **10/121 ( 8.3 )** | **1/48 ( 2.1 )** | **0.183** |
| **Malignancy** | **1/169 ( 0.6 )** | **1/121 ( 0.8 )** | **0/48 ( 0 )** | **1.000** |
| **Cerebrovascular**<br>**disease** | **5/169 ( 3.0 )** | **5/121 ( 4.1 )** | **0/48 ( 0 )** | **0.323** |
| **COPD or chronic**<br>**bronchitis** | **4/169 ( 2.4 )** | **4/121 ( 3.3 )** | **0/48 ( 0 )** | **0.579** |
| **Others** | **10/169 ( 5.9 )** | **9/121 ( 7.4 )** | **1/48 ( 2.1 )** | **0.285** |
| **Laboratory findings-**<br>**median (IQR)** |  |  |  |  |
| **SpO<sub>2</sub>-%** | **95.73** |  |  |  |
|  | **( 70–100 )** | **95.72 ( 70–100 )** | **96 ( 87–100 )** | **0.364** |
| **Total protein - g/L** | **68.69** |  |  |  |
|  | **( 36.7–98.0 )** | **66.68 ( 36.7–98.0 )** | **73.62 ( 54.7–86.3 )** | **<0.001** |
| **Albumin - g/L** | **38.8( 18–53.8 )** | **37.78 ( 18–53.8 )** | **41.19 ( 24.6–53.0 )** | **0.004** |
| **Globulin - g/L** | **29.86** |  |  |  |
|  | **( 9.8–54.5 )** | **28.87 ( 9.8–47.2 )** | **32.38 ( 23.0–54.5 )** | **0.002** |
| **Total bilirubin -**<br>**μmol/L** | **12.72** |  |  |  |
|  | **( 2.7–62.0 )** | **14.13 ( 5.8–49.2 )** | **9.88 ( 2.7–62.0 )** | **0.011** |
| **ALT-U/L** | **44.62** | **49.98 ( 2.0–547.7 )** | **30.46 ( 18.0–65.2 )** | **0.006** |
|  | ( 2.0–574.7 ) |  |  |  |
| AST-U/L | 42.83 |  |  |  |
|  | ( 5.0–584.0 ) | 47.79 ( 9.0–584 ) | 29.74 ( 5.0–72.8 ) | 0.007 |
| GGT-U/L | 78.24 ( 7–906 ) | 74.69 ( 7–906 ) | 130.3 ( 9–601 ) | 0.585 |
| Creatinine - µmol/L | 73.4 |  |  |  |
|  | ( 34.5–359 ) | 75.88 ( 39–359 ) | 68.2 ( 34.5–245 ) | 0.276 |
| White blood cell count<br>- ×10<sup>9</sup>/L | 7.07( 1.01–65 ) | 7.18 ( 1.01–65 ) | 6.77 ( 2.3–19.4 ) | 0.731 |
| Neutrophilcount -<br>×10<sup>9</sup>/L | 5.12 |  |  |  |
|  | ( 0.52–55.6 ) | 5.17 ( 0.52–55.6 ) | 5 ( 0.8–17.6 ) | 0.862 |
| lymphocyte cell count<br>- ×10<sup>9</sup>/L | 1.24 |  |  |  |
|  | ( 0.18–4.11 ) | 1.26 ( 0.18–4.11 ) | 1.18 ( 0.20–2.40 ) | 0.454 |
| Hemoglobin - g/L | 130.51 |  |  |  |
|  | ( 84–230 ) | 130.39 ( 84–230 ) | 131.13 ( 96–219 ) | 0.873 |
| Platelet count - ×10<sup>9</sup>/L | 222.35 |  |  |  |
|  | ( 55–564 ) | 219.12 ( 55–538 ) | 238.78 ( 119–564 ) | 0.328 |
| C-reactive protein -<br>mg/L | 33.77 | 31.19 |  |  |
|  | ( 0.04–285 ) | ( 0.04–254.35 ) | 40.05 ( 0.5–285 ) | 0.316 |
| PCT - µg/mL | 0.35 |  |  |  |
|  | ( 0.02–6.02 ) | 0.37 ( 0.02–6.02 ) | 0.29 ( 0.02–2.94 ) | 0.468 |
| LDH - U/L | 263.16 |  |  |  |
|  | ( 120–808 ) | 281.56 ( 120–808 ) | 227.65 ( 151–610 ) | 0.009 |
| Creatine kinase - U/L | 107.53 |  |  |  |
|  | ( 12–1381 ) | 110.94 ( 28–1381 ) | 101.81 ( 12–355 ) | 0.745 |
| CK-MB - U/L | 13.91 ( 1–73 ) | 13.02 ( 4–45.7 ) | 15.12 ( 1–73 ) | 0.305 |
| PT-s | 14.65 |  |  |  |
|  | ( 9.7–100 ) | 15.43 ( 9.7–100 ) | 13.09 ( 10.5–17 ) | 0.081 |
| D-dimer - µg/ml | 2.16 ( 0–89 ) | 2.7 ( 0–89 ) | 0.76 ( 0.02–20.58 ) | 0.292 |
| Treatment- no./total |  |  |  |  |
| no. (%) |  |  |  |  |
| Antiviral | 91/169( 53.85 ) | 73 ( 60.30 ) | 18 ( 37.50 ) | 0.007 |
| Antibiotics | 154/169 | 108 ( 89.30 ) | 46 ( 95.80 ) | 0.291 |
|  | ( 91.10 ) |  |  |  |
| Corticosteroids | 48/169( 28.40 ) | 42 ( 34.70 ) | 6 ( 12.50 ) | 0.004 |
| Outcome- no./total no. |  |  |  | 0.561 |
| (%) |  |  |  |  |
| Improvement | 165/169 | 117/121 ( 96.70 ) | 48/48 ( 100 ) |  |
|  | ( 97.60 ) |  |  |  |
| Death | 4/169 ( 2.40 ) | 4/121 ( 3.30 ) | 0/48 ( 0 ) |  |

Among the 4 age groups into which patients were stratified, groups C and D had higher proportions of severe and critical disease than the other 2 groups, and group C had the highest proportion of severe disease among the 4 groups. Group A had the least proportion of patients with dyspnea, while group C had the highest. In both group C and group D, oxygen saturation(SpO2)was lower than 94%, and group C had the lowest SpO2.The lymphocyte count was lower in groups C and D than in groups A and B. LDH in group A was the lowest. CRP, pro-calcitonin (PCT), and interleukin (IL)-6 were significantly higher in groups C and D than in groups A and B. (Table2).

**Table 2:** Clinical features of study subjects according to age group

|  | Age –yrs |  |  |  | p value |
| --- | --- | --- | --- | --- | --- |
|  | Group A 18–44 (n=52) | Group B 45–64 (n=70) | Group C 65–74 (n=35) | Group D ≥75 (n=12) |  |
| **Sex - no./total no.(%)** |  |  |  |  | **0.743** |
| Male | 21/52 ( 40.4 ) | 35/70 ( 50.0 ) | 17/35 ( 48.6 ) | 6/12 ( 50.0 ) |  |
| Female | 31/52 ( 59.6 ) | 35/70 ( 50.0 ) | 18/35 ( 51.4 ) | 6/12 ( 50.0 ) |  |
| **Disease severity-**<br>**no./total no.(%)** |  |  |  |  | **0.002** |
| Mild | 8/52 ( 15.4 ) | 3/70 ( 4.2 ) | 1/35 ( 2.9 ) | 0/12 ( 0 ) | 0.077 |
| Moderate | 41/52 ( 78.8 ) | 63/70 ( 90 ) | 24/35 ( 68.6 ) | 9/12(75.0) | 0.054 |
| Severe | 3/52 ( 5.8 ) | 2/70 ( 2.9 ) | 7/35 ( 20.0 ) <sup>AC</sup> | 1/12(8.3) | 0.015 |
| Critical | 0/52 ( 0 ) | 2/70 ( 2.9 ) | 3/35 ( 8.5 ) | 2/12(16.7)<sup>AD,BD</sup> | 0.020 |
| **No. of days from illness**<br>**onset to admission –**<br>**median(IQR)** | 8.2 ( 0–32 ) | 14.4 ( 1–40 ) <sup>AB</sup> | 12.3 ( 1–45 ) <sup>AC</sup> | 9.5 ( 0–20 ) | **0.009** |
| **Symptoms - no./total**<br>**no.(%)** |  |  |  |  |  |
| Fever | 34/52 ( 65.4 ) | 60/70 ( 85.7 ) | 27/35 ( 77.1 ) | 9/12 ( 75.0 ) | 0.073 |
| Dry cough | 33/52 ( 63.5 ) | 46/70 ( 65.7 ) | 26/35 ( 74.3 ) | 10/12 ( 83.3 ) | 0.459 |
| **Sputum production** | **10/52 ( 19.2 )** | **22/70 ( 31.9 )** | **11/35 ( 31.4 )** | **3/12 ( 25 )** | **0.43** |
| **Shortness of breath** | **3/52 ( 5.8 )** | **14/70 ( 20.0 ) <sup>AB</sup>** | **17/35 ( 48.6 ) <sup>AC</sup>** | **4/12 ( 33.3 ) <sup>AD</sup>** | **<0.001** |
| **Myalgia** | **2/52 ( 3.8 )** | **8/70 ( 22.4 )** | **4/35 ( 11.4 )** | **1/12 ( 8.3 )** | **0.395** |
| **Digestive symptoms** | **9/52 ( 17.3 )** | **11/70 ( 15.7 )** | **7/35 ( 20 )** | **2/12 ( 16.7 )** | **0.944** |
| **Comorbidities - no./total**<br>**no.(%)** |  |  |  |  |  |
| **Any comorbidity** | **6/52 ( 11.5 )** | **21/70 ( 30 )** | **19/35 ( 54.3 )** | **6/12 ( 50.0 )** | **<0.001** |
| **Hypertension** | **3/52 ( 5.8 )** | **10/70 ( 14.3 )** | **12/35 ( 34.3 )** | **5/12 ( 41.7 )** | **0.001** |
| **Cardiovascular disease** | **2/52 ( 3.8 )** | **7/70 ( 10.0 )** | **5/35 ( 14.3 )** | **0/12 ( 0 )** | **0.257** |
| **Diabetes** | **0/52 ( 0 )** | **2/70 ( 2.9 )** | **8/35 ( 22.9 )** | **1/12 ( 8.3 )** | **<0.001** |
| **Malignancy** | **0/52 ( 0 )** | **0/70 ( 0 )** | **1/35 ( 2.9 )** | **0/12 ( 0 )** | **0.278** |
| **Cerebrovascular disease** | **0/52 ( 0 )** | **0/70 ( 0 )** | **1/35 ( 2.9 )** | **4/12 ( 33.3 )** | **<0.001** |
| **COPD** | **0/52 ( 0 )** | **0/70 ( 0 )** | **3/35 ( 8.5 )** | **1/12 ( 8.3 )** | **0.006** |
| **Others** | **2/52 ( 3.8 )** | **5/70 ( 7.1 )** | **2/35 ( 5.7 )** | **1/12 ( 8.3 )** | **0.825** |
| **Laboratory findings-**<br>**median (IQR)** |  |  |  |  |  |
| SpO<sub>2</sub>-% | 97.31 ( 93–100 ) | 96.17 ( 70–99 ) | 93.26 ( 76–98 ) | 93.55 ( 80–98 ) | 0.032 |
| Total protein - g/L | 73 ( 57–86.3 ) | 68.5 ( 36.7–98 )<sup>AB</sup> | 63.5 ( 53.4–82.2 )<sup>AC,BC</sup> | 65.7 ( 51–81.8 )<sup>AD</sup> | <0.001 |
| Albumin - g/L | 43.3 ( 32.6–53 ) | 38.6 ( 18–53.8 ) | 33.3 ( 20–44.5 ) | 36.1 ( 27–44 ) | <0.001 |
| Globulin - g/L | 29.7 ( 22–39.7 ) | 29.9 ( 9.8–47.2 ) | 30 ( 19–54.5 ) | 29.5 ( 18–41.8 ) | 0.947 |
| Total bilirubin - μmol/L | 11.9 ( 2.91–49.2 ) | 11.5 ( 2.8–33 ) | 12.8 ( 2.7–48 ) | 20.5 ( 5.9–62 ) | 0.151 |
| ALT-U/L | 45 ( 7–547.7 ) | 50.4 ( 11–479 ) | 32.3 ( 10–81 ) | 41.5 ( 2–101 ) | 0.626 |
| AST-U/L | 35.1 ( 9–352.7 ) | 49.2 ( 13–584 ) | 38.1 ( 5–157 ) | 51.6 ( 20–109.2 ) | 0.540 |
| GGT-U/L | 93.5 ( 10–906 ) | 82.2 ( 8–527 ) | 72.8 ( 7–601) | 44 ( 7–906 ) | 0.766 |
| Creatinine - μmol/L | 61 ( 34.5–91 ) | 74.2 ( 41–241 ) | 78 ( 46.1–245 ) | 104.5 ( 60–359 ) | 0.001 |
| White blood cell count -<br>×10<sup>9</sup>/L | 6.78 ( 2.3–60.9 ) | 7.46 ( 2.34–65 ) | 7.1 ( 1.01–19.4 ) | 5.9 ( 1.92–13.61 ) | 0.893 |
| Neutrophil count -<br>×10<sup>9</sup>/L | 4.76 ( 0.8–55.6 ) | 5.2 ( 1.12–20.71 ) | 5.62 ( 0.52–17.6 ) | 5.1 ( 1.03–10.66 ) | 0.91 |
| Lymphocyte count -<br>×10<sup>9</sup>/L | 1.43(0.43–3.3) | 1.31(0.23–4.11) | 0.94(0.18–2.2)<sup>AC,BC</sup> | 0.85(0.2–2.07)<sup>AD,BD</sup> | 0.001 |
| Hemoglobin - g/L | 137.47 ( 102–230 ) | 131.48 ( 100–191 ) | 123.5 ( 84–177 ) | 114.72 ( 87–147 ) | 0.001 |
| Platelet count - ×10<sup>9</sup>/L | 218.5 ( 55–431 ) | 225.73 ( 84–564 ) | 243 ( 100–538 ) | 167.36 ( 85–250 ) | 0.108 |
| C-reactive protein -<br>mg/L | 17.76 ( 0.04–78.95 ) | 30.11 ( 0.4–149.68 ) | 46.84 ( 0.4–285 ) | 88.57 ( 1–254.35 ) | 0.006 |
| PCT -µg/mL | 0.21 ( 0.02–0.67 ) | 0.34 ( 0.04–2.57 ) | 0.49 ( 0.03–6.02 ) | 0.56 ( 0.08–2.05 ) | 0.017 |
| LDH - U/L | 212.86 ( 120–681 ) | 275.74 ( 123–686 ) <sup>AB</sup> | 282.2 ( 175–544 ) <sup>AC</sup> | 369.09 ( 177–808 ) <sup>AD</sup> | <0.001 |
| Creatinekinase - U/L | 117.42 ( 31.4–1381 ) | 101.87 ( 28–355 ) | 88.87 ( 12–216 ) | 120.6 ( 43–600 ) | 0.891 |
| CK-MB - U/L | 14.09 ( 5–68 ) | 12.75 ( 4–45.7 ) | 17.34 ( 6–73 ) | 16.52 ( 5–32.26 ) | 0.405 |
| PT-s | 0.24 ( 0.01–1.23 ) | 0.88 ( 0–7.8 ) | 4.23 ( 0.02–80 ) | 10.66 ( 0.18–89 ) <sup>AD,BD,CD</sup> | <0.001 |
| D-dimer - µg/ml | 13.16 ( 9.7–15.8 ) | 13.37 ( 9.7–29.3 ) | 15.57 ( 10.9–76 ) | 21.87 ( 12.6–100 ) <sup>AD,BD</sup> | 0.001 |
| Immunology – median<br>(IQR) |  |  |  |  |  |
| IgG- g/L | 11.53 ( 7.84–15.16 ) | 13.39 ( 7.42–20.81 ) | 12.66 ( 7.96–16.69 ) | 12.75 ( 7.24–19.18 ) | 0.647 |
| IgA- g/L | 2.15 ( 1.32–3.44 ) | 2.63 ( 1.37–5.09 ) | 2.07 ( 1.47–3.34 ) | 2.65 ( 2.38–4.71 ) | 0.26 |
| IgM- g/L | 1.42 ( 0.64–2.29 ) | 1.53 ( 0.64–3.15 ) | 1.37 ( 0.82–2.55 ) | 1.18 ( 0.38–2.49 ) | 0.662 |
| IgE- g/L | 74.77 ( 2.4–480.1 ) | 124.38 ( 1.3–1100 ) | 32.85 ( 0.8–76.1 ) | 154.15 ( 2.8–1008 ) | 0.717 |
| IL-6- pg/mL | 1.69 ( 15–2.84 ) | 6.76 ( 1.5–42.53 ) | 33.57 ( 1.5–223.81 ) | 71.34 ( 2.89–252.7 ) | 0.007 |
| C3- g/L | 0.94 ( 0.8–1.3 ) | 0.94 ( 0.6–1.3 ) | 0.85 ( 0.8–1 ) | 0.94 ( 0.6–1.4 ) | 0.701 |
| **C4- g/L** | **0.199 ( 0.1–0.4 )** | **0.18 ( 0.1–0.3 )** | **0.18 ( 0.1–0.2 )** | **0.22 ( 0.1–0.4 )** | **0.61** |
| **CD3+ cell count - cell/μl** | **1057.83 ( 243–1736)** | **785.66(377–1357)** | **577.28(142–1310)** | **–** | **0.015** |
| **CD4+ cell count - cell/μl** | **666.77(79–1320)** | **520.5(207–1000)** | **402.57(79–883)** | **–** | **0.081** |
| **CD8+ cell count - cell/μl** | **391.05(107–892)** | **265.16(75–548)** | **174.71(58–427)** | **–** | **0.011** |
| **CD4+ cell percentage**<br>**- %** | **–** | **45.87 ( 42.32–50.93 )** | **39.85 ( 10.83–59.3 )** | **31.23** | **0.546** |
| **CD8+ cell percentage**<br>**- %** | **–** | **24.77 ( 22.08–28.67 )** | **26.18 ( 14.67–46.04 )** | **15.54** | **0.436** |
| **CD3+ cell percentage**<br>**- %** | **–** | **72.84 ( 70.68–74.79 )** | **68.87 ( 58.64–86.66 )** | **47.77** | **0.195** |
| **NK cell percentage - %** | **–** | **5.18 ( 3.54–6.47 )** | **11.71 ( 2.69–27.59 )** | **7.64** | **0.346** |
| **Number of lung lobes**<br>**involved - no./total**<br>**no.(%)** |  |  |  |  |  |
| **5** | **12/45 ( 26.7 )** | **29/48 ( 60.4 )** | **16/25 ( 64.0 )** | **5/10 ( 50.0 )** | **0.003** |
| **4** | **2/45 ( 4.5 )** | **7/48 ( 14.6 )** | **2/25 ( 8.0 )** | **0/10 ( 0 )** | **0.336** |
| **3** | **3/45 ( 6.7 )** | **4/48 ( 8.3 )** | **4/25 ( 16.0 )** | **4/10 ( 4.0 )** | **0.032** |
| **2** | **11/45 ( 24.4 )** | **4/48 ( 8.3 )** | **2/25 ( 8.0 )** | **0/10 ( 0 )** | **0.073** |
| **1** | **11/45 ( 24.4 )** | **2/48 ( 4.2 )** | **0/25 ( 0 )** | **1/10 ( 10.0 )** | **0.003** |
| 0 | 6/45 ( 13.3 ) | 2/48 ( 4.2 ) | 1/25 ( 4.0 ) | 0/10 ( 0 ) | 0.343 |
| **Treatment - no./total**<br>**no.(%)** |  |  |  |  |  |
| Antiviral | 21/52 ( 40.4 ) | 39/70 ( 55.7 ) | 25/35 ( 71.4 ) | 6/12 ( 50.0 ) | 0.047 |
| Antibiotics | 48/52 ( 92.3 ) | 66/70 ( 94.3 ) | 32/35 ( 91.4 ) | 8/12 ( 66.7 ) | 0.001 |
| Corticosteroids | 6/52 ( 11.5 ) | 19/70 ( 27.1 ) | 18/35 ( 51.4 ) | 0/12 ( 0 ) | 0.001 |
| **Prognosis - no./total**<br>**no.(%)** |  |  |  |  | 0.24 |
| Death | 0/52 ( 0 ) | 2/70 ( 2.9 ) | 1/35 ( 2.9 ) | 1/12 ( 8.3 ) |  |
| improve | 52/52 ( 100.0 ) | 68/70 ( 97.1 ) | 34/35 ( 97.1 ) | 11/12 ( 91.7 ) |  |

IQR=interquartile range, COPD=chronic obstructive pulmonary disease, ALT=alanine transaminase, AST=aspartate aminotransferase, GGT=gamma glutamyl transferase, PCT=procalcitonin, LDH=lactate dehydrogenase, CK-MB=creatin kinase-MB, PT=prothrombin time, Ig=immunoglobulin, IL=interleukin, C3=complement 3, C4=complement 4, CD=cluster of differentiation, NK= natural killer Comparison between groups (P<0.05):AB= A compared with B, AC= A compared with C, AD=A compared with D, BC=B compared with C, BD=B compared with D, CD=C compared with D.

Fifty-two patients (30.8%) in the study had comorbidities. Compared with patients who had no comorbidities, the percentage of critical COVID-19 was higher in the comorbidities group(11.6%vs.0.9%; p<0.05).Patients with comorbidities were also more likely to have dyspnea(36.5% vs. 16.2%; p<0.05).The total protein and albumin values and lymphocyte counts in patients with comorbidities were lower, while increase in CRP and LDH was more obvious in this group.All4 patients who died had comorbidities(Table3).

**Table 3.** Features of study subjects based on presence of comorbidities

|  | **Have comorbidities** | **Have no comorbidities** | **p value** |
| --- | --- | --- | --- |
|  | **( N=52 )** | **( N=117 )** |  |
| **Disease severity-**<br>**no./total no.(%)** |  |  | **0.004** |
| **Mild** | **1/52 ( 1.9 )** | **11/117 ( 9.4 )** | **0.108** |
| **Moderate** | **40/52 ( 76.9 )** | **97/117 ( 82.9 )** | **0.360** |
| **Severe** | **5/52 ( 9.6 )** | **8/117 ( 6.8 )** | **0.542** |
| **Critical** | **6/52 ( 11.6 )** | **1/117 ( 0.9 )** | **0.004** |
| **Sex - no./total no.(%)** |  |  | **0.368** |
| **Male** | **27/52 ( 51.9 )** | **52/117 ( 44.4 )** | **-** |
| **Female** | **25/52 ( 48.1 )** | **65/117 ( 55.6 )** | **-** |
| **Age in yrs- median**<br>**(IQR)** | **59.88 ( 25–86 )** | **48.69 ( 18–92 )** | **0.001** |
| **Days from onset of**<br>**illness to confirmation**<br>**of diagnosis -**<br>**median (IQR)** | **11.18 ( 0–40 )** | **11.86 ( 0–45 )** | **0.691** |
| **Symptoms - no./total**<br>**no.(%)** |  |  |  |
| **Fever** | **43/52 ( 82.7 )** | **87/117 ( 74.4 )** | **0.235** |
| **Dry cough** | **39/52 ( 75.0 )** | **76/117 ( 68.0 )** | **0.196** |
| **Sputum production** | **17/52 ( 33.3 )** | **29/117 ( 24.8 )** | **0.253** |
| **Shortness of breath** | **19/52 ( 36.5 )** | **19/117 ( 16.2 )** | **0.004** |
| **Digestive symptoms** | **6/52 ( 11.5 )** | **9/117 ( 7.7 )** | **0.604** |
| **Myalgia** | **9/52 ( 17.6 )** | **20/117 ( 17.1 )** | **0.931** |
| **Laboratory**<br>**findings-median (IQR)** |  |  |  |
| **SpO<sub>2</sub>- %** | **94.4 ( 70–99 )** | **96.3 ( 72–100 )** | **0.121** |
| Total protein - g/L | 65.66 ( 39–84 ) | 70.06 ( 36.7–98 ) | 0.004 |
| Albumin - g/L | 36.34 ( 18–53 ) | 39.9 ( 20–53.8 ) | 0.002 |
| Globulin - g/L | 29.28 ( 18–47.2 ) | 30.12 ( 9.8–54.5 ) | 0.458 |
| Total bilirubin -<br>μmol/L | 16.65 ( 4.3–62 ) | 10.91 ( 2.7–49.2 ) | 0.006 |
| ALT-U/L | 33.96 ( 2–120 ) | 49.32 ( 10–574.7 ) | 0.046 |
| AST-U/L | 38.94 ( 5–207 ) | 44.54 ( 9–584 ) | 0.476 |
| GGT-U/L | 71.33 ( 10–601 ) | 81.99 ( 7–906 ) | 0.706 |
| Creatinine - μmol/L | 81.69 ( 43–359 ) | 69.45 ( 34.5–241 ) | 0.177 |
| White blood cell count<br>- ×10<sup>9</sup>/L | 7.74 ( 1.01–65 ) | 6.77 ( 1.92–60.9 ) | 0.404 |
| Neutrophil count -<br>×10<sup>9</sup>/L | 5.44 ( 0.52–20.71 ) | 4.96 ( 0.8–55.6 ) | 0.646 |
| Lymphocyte count -<br>×10<sup>9</sup>/L | 1.07 ( 0.18–2.66 ) | 1.31 ( 0.26–4.11 ) | 0.032 |
| Hemoglobin - g/L | 127.95 ( 84–191 ) | 131.57 ( 100–230 ) | 0.331 |
| Platelet count - ×10<sup>9</sup>/L | 213.34 ( 85–392 ) | 226.73 ( 55–564 ) | 0.292 |
| C-reactive protein -<br>mg/L | 49.72 ( 0.3–254.35 ) | 27.22 ( 0.04–285 ) | 0.026 |
| PCT -μg/mL | 0.53 ( 0.02–6.02 ) | 0.27 ( 0.02–1.74 ) | 0.091 |
| ESR - mm/h | 44.67 ( 20–97.3 ) | 37.62 ( 9–119 ) | 0.175 |
| LDH - U/L | 327.21 ( 123–808 ) | 235.51 ( 120–681 ) | 0.002 |
| Creatinekinase - U/L | 106.45 ( 12–600 ) | 107.91 ( 28–1381 ) | 0.961 |
| CK-MB - U/L | 16.37 ( 5–73 ) | 12.68 ( 1–45.7 ) | 0.181 |
| D-dimer -μg/ml | 3.33 ( 0–80 ) | 0.71 ( 0.01–8.00 ) | 0.175 |
| PT-s | 18.18 ( 10.9–100 ) | 13.22 ( 9.7–17.4 ) | 0.101 |
| Immunological<br>findings- median (IQR) |  |  |  |
| IgG- g/L | 2.32 ( 1.37–3.54 ) | 2.46 ( 1.32–5.09 ) | 0.625 |
| IgA- g/L | 1.43 ( 0.46–2.98 ) | 1.37 ( 0.38–3.15 ) | 0.785 |
| IgM- g/L | 120.19 ( 0.8–1100 ) | 99.05 ( 1.3–1008 ) | 0.770 |
| IgE- g/L | 11.5 ( 7.24–16.88 ) | 13.03 ( 7.42–20.81 ) | 0.229 |
| C3- g/L | 0.91 ( 0.6–1.2 ) | 0.93 ( 0.6–1.4 ) | 0.719 |
| C4- g/L | 0.18 ( 0.1–0.3 ) | 0.2 ( 0.1–0.4 ) | 0.433 |
| IL-6- pg/mL | 17.67 ( 1.5–124.8 ) | 23.42 ( 0–252.7 ) | 0.773 |
| CD3+ cell count -<br>cell/μl | 65.6 ( 58.64–75.45 ) | 69.59 ( 47.77–86.66 ) | 0.487 |
| CD4+ cell count -<br>cell/μl | 12.48 ( 2.69–27.59 ) | 8.9 ( 3.54–18.76 ) | 0.379 |
| CD8+ cell counts -<br>cell/μl | 14.99 ( 1.8–37.85 ) | 17.61 ( 3.3–37.33 ) | 0.673 |
| CD4+ cell percentage<br>- % | 36.61 ( 10.83–59.3 ) | 42.41 ( 31.23–55 ) | 0.376 |
| CD8+ cell percentage<br>- % | 26.85 ( 14.67–46.04 ) | 28.8 ( 15.54–73.05 ) | 0.816 |
| Days before negative<br>nucleic acid<br>test-median (IQR) | 8.18 ( 2–30 ) | 6.16 ( 1–18 ) | 0.092 |
| Treatment - no./total<br>no. (%) |  |  |  |
| Antibiotics | 33/52 ( 63.50 ) | 58/117 ( 49.6 ) | 0.095 |
| Antiviral | 49/52 ( 94.2 ) | 105/117 ( 89.7 ) | 0.558 |
| Corticosteroids | 16/52 ( 30.8 ) | 32/117 ( 27.4 ) | 0.649 |
| Outcome - no./total no.<br>(%) |  |  | 0.008 |
| Death | 4/52 ( 7.7 ) | 0/117 ( 0 ) |  |
| Improvement | 48/52 ( 92.3 ) | 117/117 ( 100 ) |  |

IQR=interquartile range, ALT=alanine transaminase, AST=aspartate aminotransferase, GGT=gamma glutamyl transferase, PCT=procalcitonin, LDH=lactate dehydrogenase, CK-MB=creatine kinase-MB, ESR=erythrocyte sedimentation rate, PT=prothrombin time, Ig=immunoglobulin

To determine the association of disease severity with age, we divided all patients into 5 groups at intervals of 15 years, and analyzed the frequencies of severe and critical disease. We found that older patients had higher proportion of severe or critical disease(Table 4).The 169 patients were also divided into two groups—mild group(including mild type and moderate type COVID-19,n=148) and severe group(including severe and critical type COVID-19, n=21)—and we then analyzed the risk factors for disease severity. The results showed that lymphocyte count and SpO2 at admission were factors that determined the risk factor of disease severity (Table5).

**Table 4:** Relationship between age and disease severity(N=169)

| Variable | B | Wald$\chi^2$ | P | 95%CI |
| --- | --- | --- | --- | --- |
| Q1 | -4.604 | 32.983 | 0.000 | ( -6.175~-3.033 ) |
| Q2 | 0.512 | 0.586 | 0.444 | ( -0.799~1.823 ) |
| Q3 | 1.770 | 6.063 | 0.014 | ( 0.361~3.180 ) |
| Age group<br>(control: group 5) |  |  |  |  |
| 1 | -2.675 | 8.098 | 0.004 | ( -4.518~-0.883 ) |
| 2 | -2.382 | 8.146 | 0.004 | ( -4.017~-0.746 ) |
| 3 | -1.768 | 5.022 | 0.025 | ( -3.314~-0.222 ) |
| 4 | -0.628 | 0.691 | 0.406 | ( -2.110~0.853 ) |

Q1=Mild disease, Q2=Moderate disease, Q3=Severe disease, Q4=Critical disease
Age group (years, 15-year intervals): 1=18–32, 2=33–47, 3=48–62, 4=63–77, 5=78–92; B=regression coefficient; OR=odds ratio; 95%CI=95% confidence interval

**Table 5:**
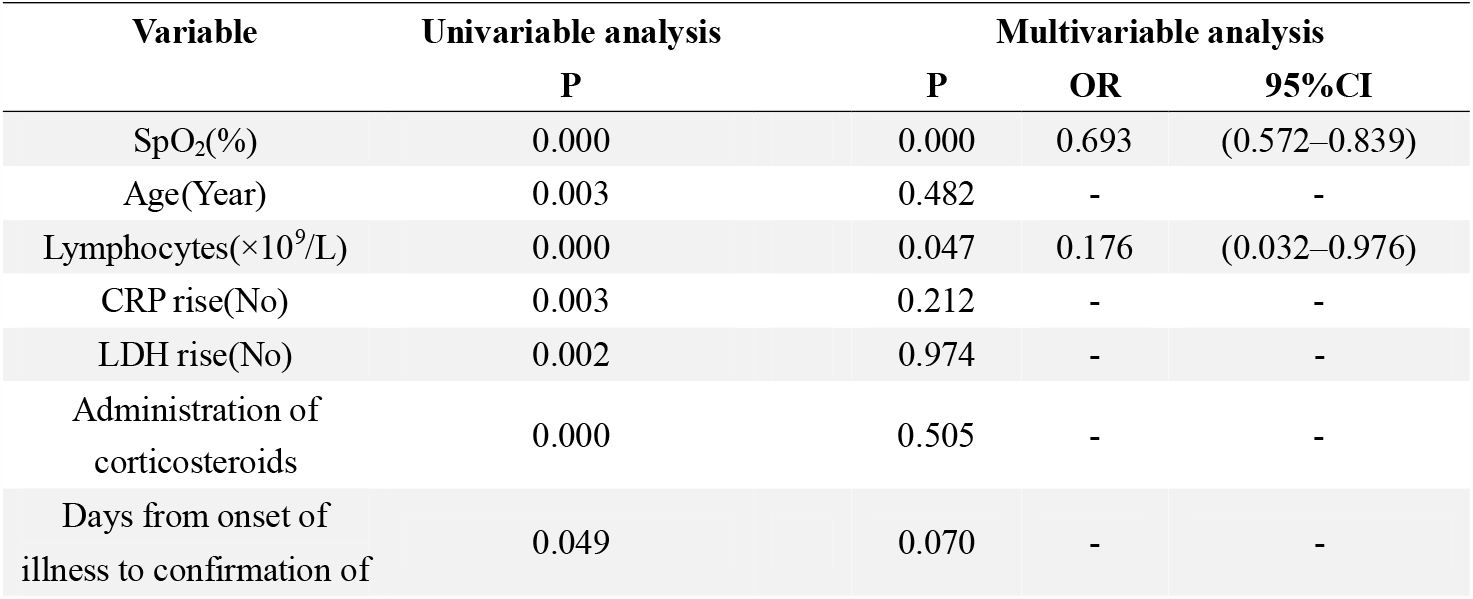
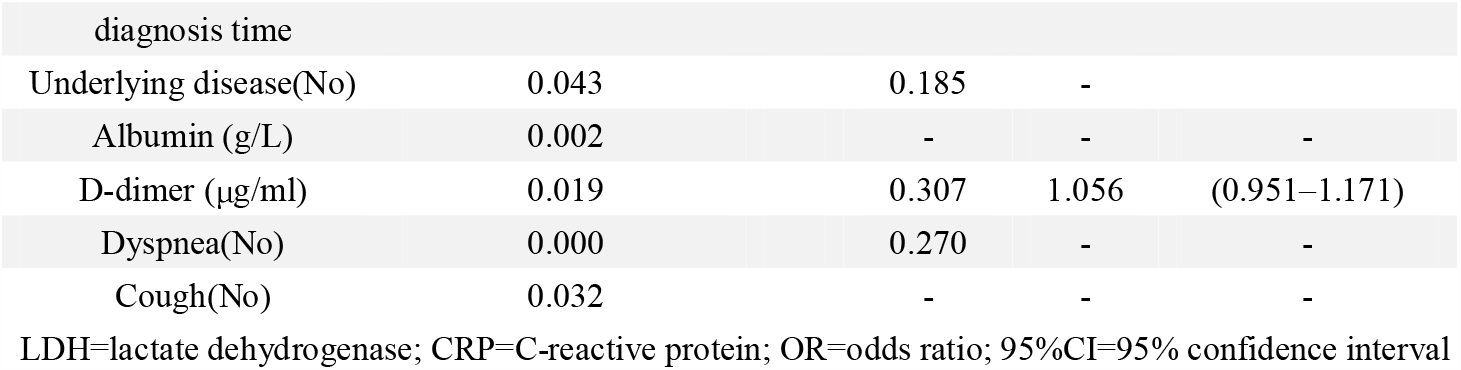
Univariable and multivariable analyses of risk factors for disease severity(N=169)

A total of 116 patients with CT image data were enrolled in this study. The average time from onset of illness to CT imaging was 10.73±7.97(0–34)days. The first CT imaging findings of the 116 patients were analyzed, and we found that 84(72.41%) patients had bilateral lung involvement,88(75.86%) patients hadperipheral lesions, and99(85.3%) patients had involvement of two or more lung lobes. The most common CT features were consolidation (66.4%), GGO (55.2%), linear opacity (55.20%), and mixed GGO and consolidation (33.60%). Forty (34.48%) patients had vascular thickening.

We further divided the patients into 6 groups according to the number of days from onset of illness to repeat chest CT imaging: group1(≤7days, n=59), group2(8–14days, n=92), group3(15–21days, n=109), group4(22–28days, n=60), group5(29–45days, n=52), and group6(46–60days, n=18). Among the groups, group 1 had the highest percentage of GGO (48cases,81.4%), and the least percentage of linear opacity (20cases, 33.9%). Group 2 had the highest percentage of consolidation (72cases,78.3%). The proportion of cases with linear opacity gradually increased from group1 to group6, reaching 98.1% in group6.There were 4casesof reticular changes in group1,and the proportion gradually increased to group 6, in which there were 7cases(Table 6).

**Table 6:**
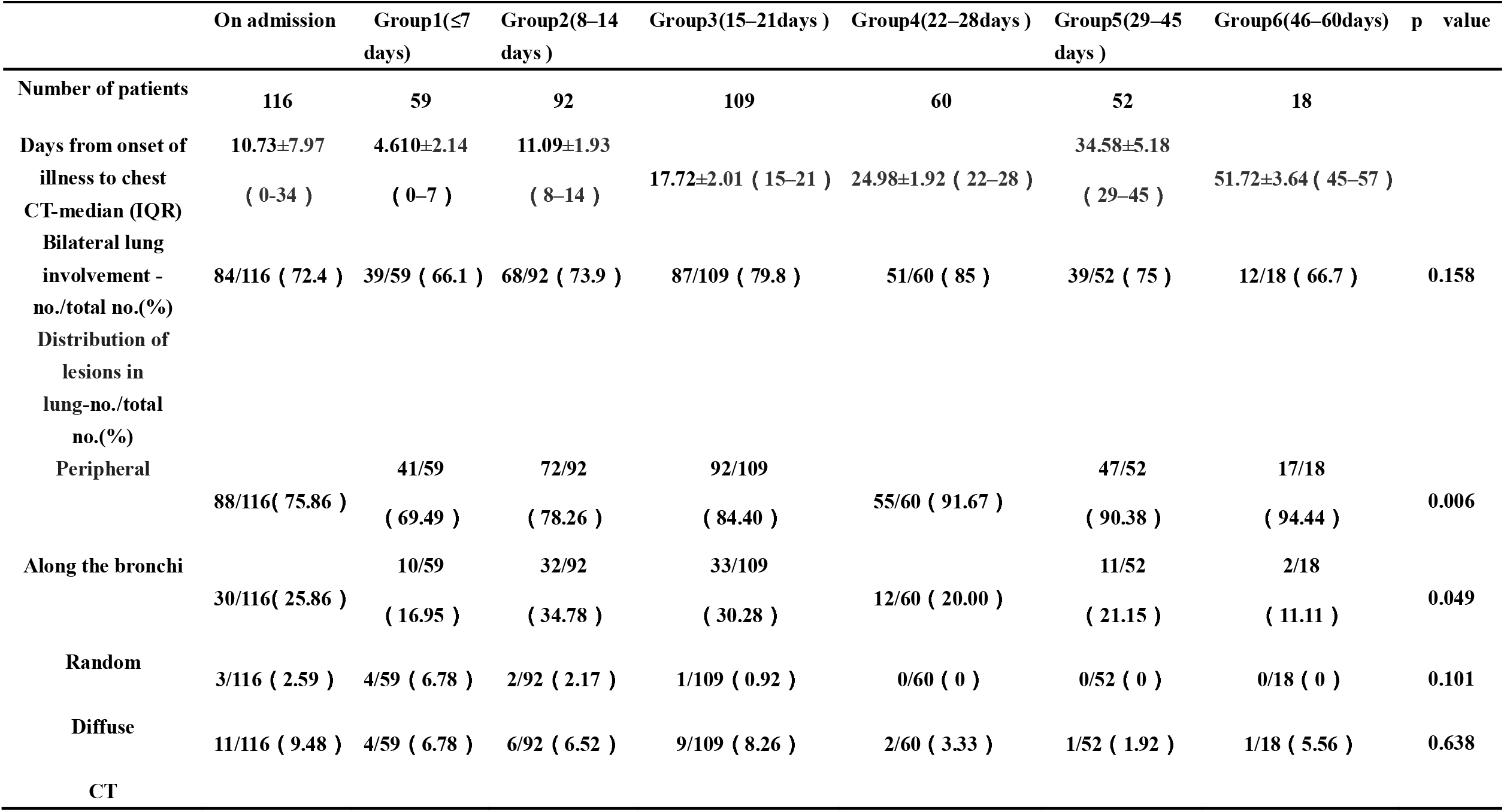
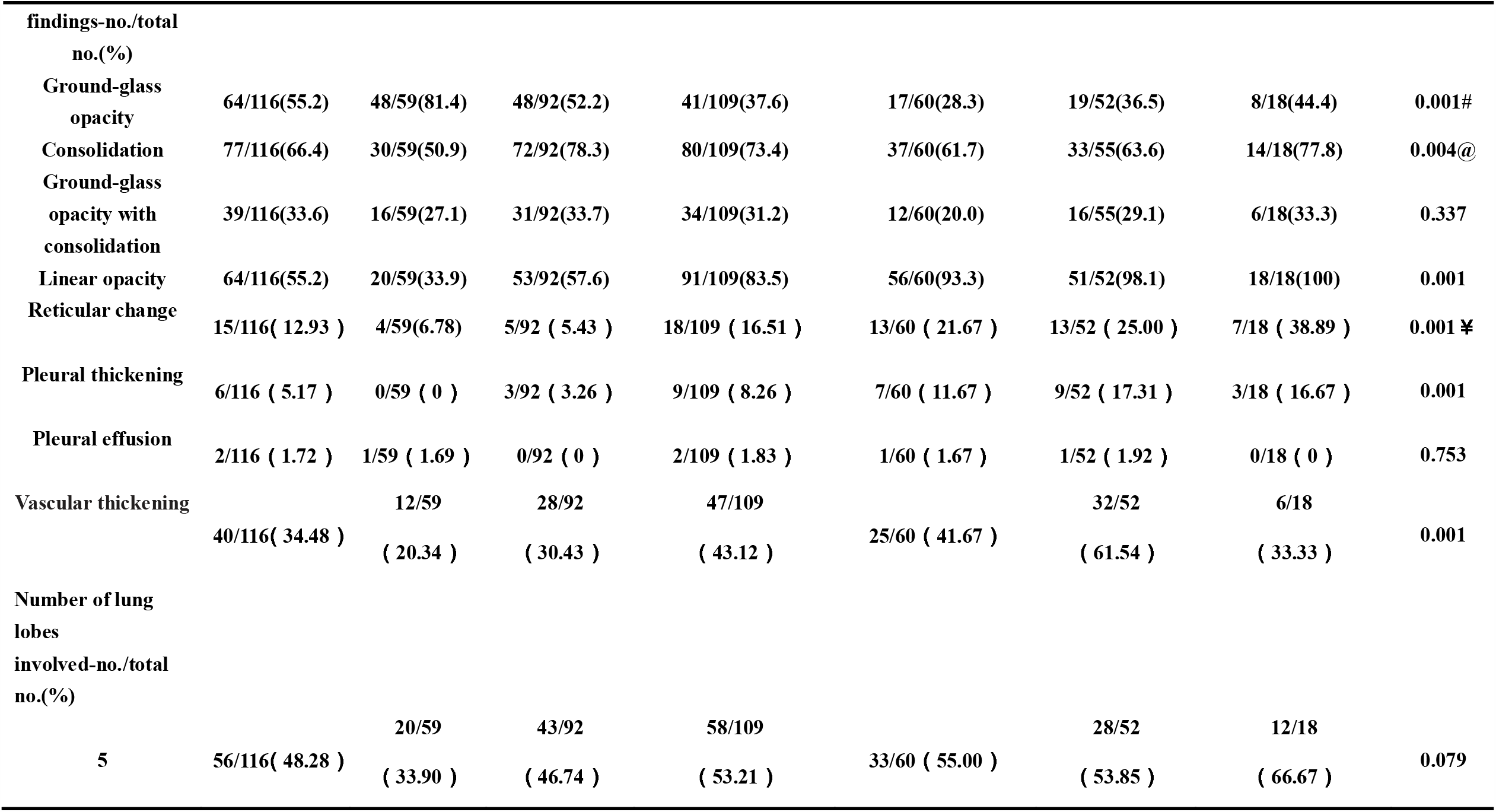
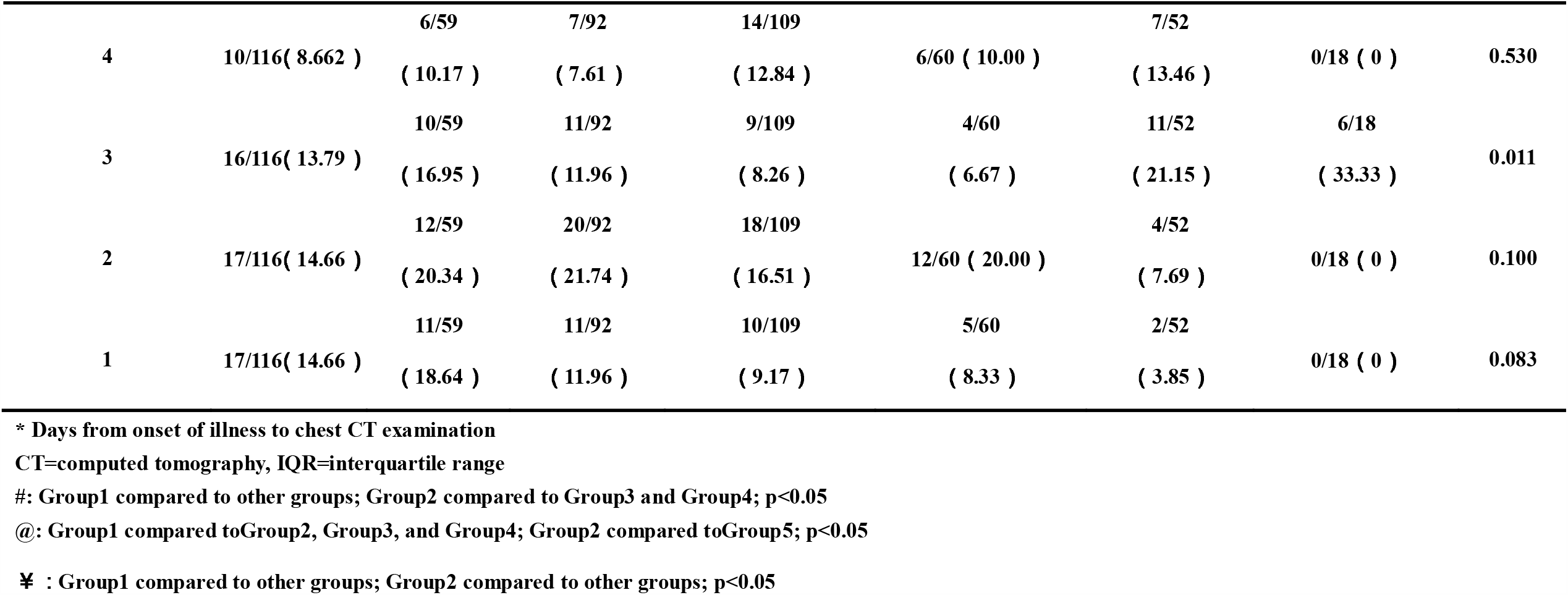
Chest CT findings of study subjects on different days*

To monitor the changes in CT images during the course of the disease, we considered the dynamic changes in the CT images of two patients from Guangxi, from onset to improvement of the disease: one with severe disease (patient1, Figure1) and the other with moderate disease (patient 2, Figure 2). As shown in Figure 1, patient 1 had ground glass opacity on chest CT in the early stage of the disease, and consolidation was noted on chest CT done on day 13 from onset of illness, as the patient’s clinical symptoms improved. Consolidation had resolved on day 18 from onset of illness. Finally, the patient had linear opacity on day 23 from onset of illness. There were fifty-one patients when their clinical symptoms improved and they had checked chest CT (no more than 1 day before and after). Among them, the first CT findings were mainly GGO in 36 cases. When their clinical symptoms improved, 16 cases (44.4%) showed resolution of ground glass opacities but increased consolidation on repeat CT imaging, while16 cases (44.4%) showed both GGO and resolution of consolidation.

**Figure 1:**
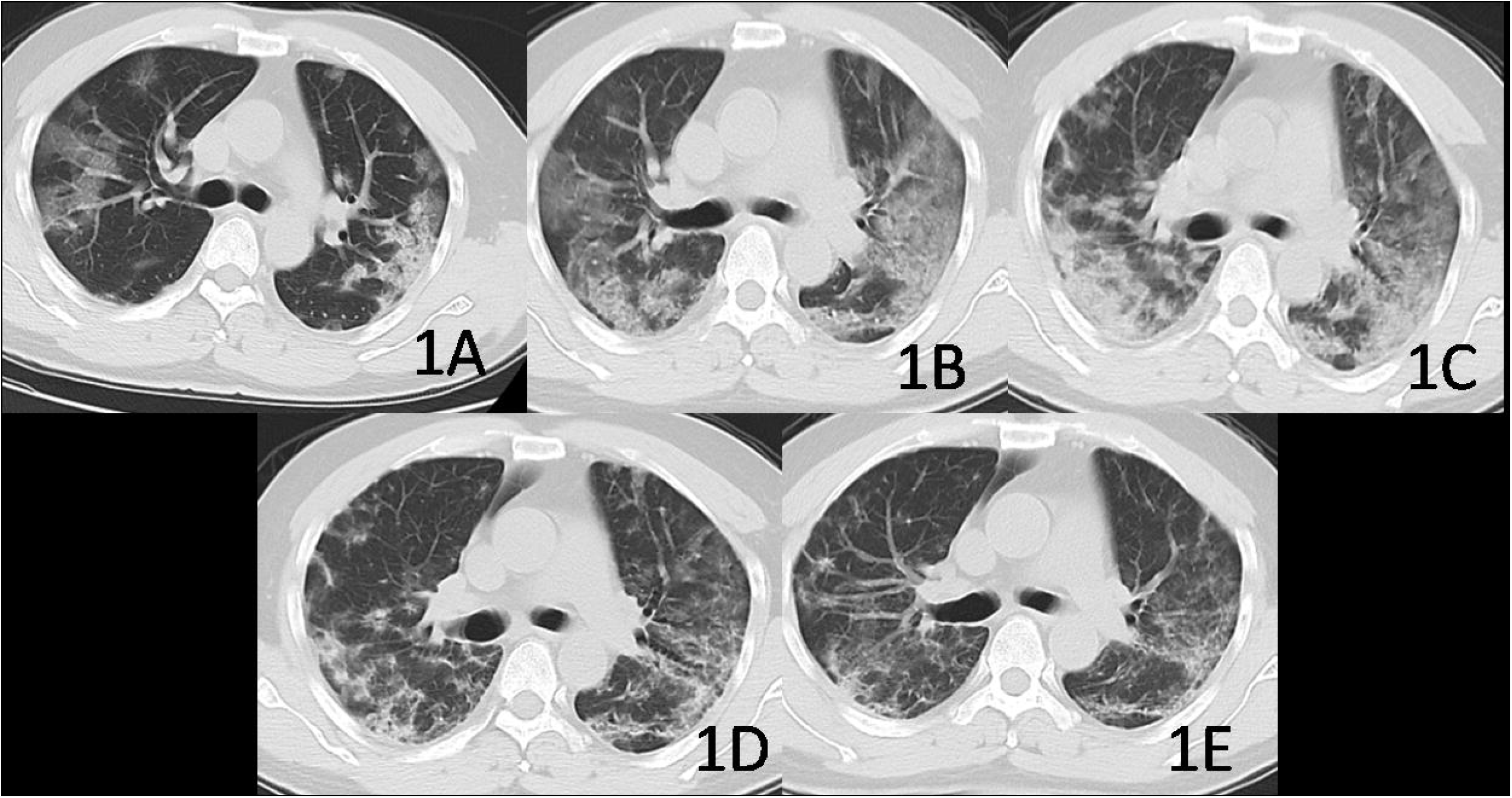
Computed tomography findings in the lungs on different days in the course of the disease in a patient with severe disease.(a)4 days from onset of illness.(b)8 days from illness onset(patient’s clinical symptoms began to improve).(c)13 days from onset of illness. (d)18 days from onset of illness. (e)23 days from onset of illness onset.

**Figure 2:**
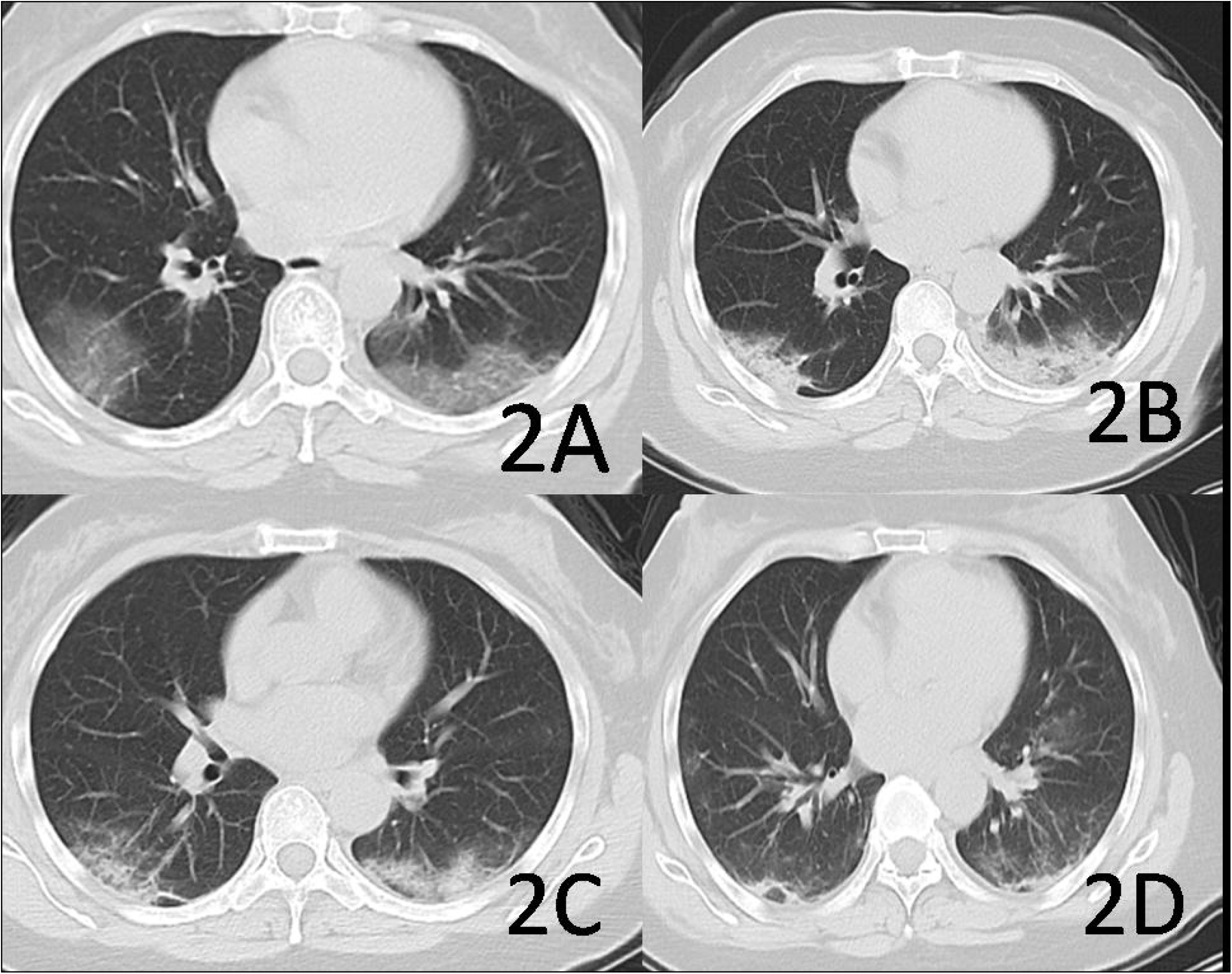
Computed tomography findings in the lungs on different days in the course of the disease in a patient with moderate disease. (a)5 days from onset of illness. (b)10 days from onset of illness (patient’s clinical symptoms began to improve). (c)15 days from onset of illness. (d)21 days from onset of illness.

All 169 patients had positive real-time RT-PCR for COVID-19 nucleic acid on nasopharyngeal swab specimens. Twenty-five patients underwent nucleic acid test on stool samples: seven of them were positive, and six among them had diarrhea. Twenty patients underwent nucleic acid test of urine samples: one was positive, but he did not have symptoms of urinary tract infection. Vaginal secretions and semen tests were performed in 6 patients, all of which were negative. We designed three prognostic models: model 1 was based on number of days from positive COVID-19 nucleic acid test to negative test, model 2 was based on number of days from onset of illness to improvement in symptoms, and model 3 was based on whether the second CT scan was better than the first. There were 79 cases included in model 1, and multivariable analysis showed that lymphocyte count was significantly associated with the number of independent risk factor of days for positiveCOVID-19 nucleic acid test to become negative(Table 7, Figure 3).There were 53 cases included in model 2, and multivariable analysis showed that number of days from onset of illness to confirmation of diagnosis was significantly associated with an independent risk factor of the time it took for symptoms to improve (Table 8, Figure 4).The average number of days from the first CT scan to the second CT scan was 5.35 days, and we took 6 days as the time point for evaluating whether the CT scans improved or not. A total of 62 patients were included in model 3, and multivariable analysis showed that number of days from onset of illness to confirmation of diagnosis and disease severity were significantly associated with independent risk factors of chest CT improvement (Table 9).

**Table 7:** Univariable and multivariable analysis of factors associated with the time it takes for nucleic acid test to turn negative (N=79)

| Variable | Univariable analysis | Multivariable analysis |  |  |
| --- | --- | --- | --- | --- |
|  | P | P | B | 95%CI |
| Sex(Male) | 0.079 | - | - | - |
| Age(Year) | 0.278 | - | - | - |
| Lymphocytes($\times 10^9/L$) | 0.005 | 0.014 | -2.974 | (-5.327~-0.620) |
| CRP(mg/L) | 0.021 | 0.954 | 0.001 | (-0.022~0.024) |
| LDH(U/L) | 0.005 | 0.730 | 0.003 | (-0.012~0.017) |
| Corticosteroids | 0.005 | 0.137 | -1.710 | (-3.978~0.557) |
| Antibiotics | 0.024 | 0.154 | -1.563 | (-3.723~0.598) |

LDH=lactate dehydrogenase; CRP=C-reactive protein; B=Regression coefficients; 95%CI=95% confidence interval

**Table 8:** Univariable and multivariable analysis of factors associated with the time it takes for symptoms to improve(N=53)

| Variable | Univariable analysis | Multivariable analysis |  |  |
| --- | --- | --- | --- | --- |
|  | P | P | B | 95%CI |
| Sex(Male) | 0.724 | - | - | - |
| Age(Year) | 0.052 | 0.095 | 0.146 | (-0.025~0.319) |
| Lymphocytes($\times 10^9/L$) | 0.029 | 0.440 | 1.713 | (-2.708~6.134) |
| CRP(mg/L) | 0.993 | - | - | - |
| LDH($\mu/L$) | 0.792 | - | - | - |
| Administration of corticosteroid | 0.323 | - | - | - |
| Timefrom onset of illness to confirmation of diagnosis (days) | 0.000 | 0.001 | 0.527 | (0.219~0.835) |

LDH=lactate dehydrogenase; CRP=C-reactive protein; B=Regression coefficients; 95%CI=95% confidence interval

**Table 9:** Univariable and multivariable analysis of factors associated with chest CT improvemen(t N=62)

| Variable | Univariable analysis | Multivariable analysis |  |  |
| --- | --- | --- | --- | --- |
|  | P | P | HR | 95%CI |
| Sex(Male) | 0.023 | 0.109 | 0.608 | (0.331–1.117) |
| Age(Year) | 0.195 | - | - | - |
| Lymphocytes($\times 10^9/L$) | 0.088 | 0.278 | 1.374 | (0.774–2.440) |
| CRP(mg/L) | 0.561 | - | - | - |
| LDH(U/L) | 0.899 | - | - | - |
| Administration of corticosteroid | 0.540 | - | - | - |
| Time from onset of illness to confirmation of diagnosis (days) | 0.075 | 0.013 | 1.019 | (1.019–1.168) |
| Disease severity | 0.045 | 0.061 | - | - |
| Moderate(compared to mild mild) | - | 0.008 | 0.261 | (0.097–0.704) |
| Severe(compared to mild mild) | - | 0.057 | 0.192 | (0.035–1.049) |
| Critical(compared to mild mild) | - | 0.971 | 0 | 0 |
| Time of onset to nucleic acid positive time(days) | 0.059 | 0.853 | 1.009 | (0.918–1.110) |

CT=computed tomography, LDH=lactate dehydrogenase, CRP=C-reactive protein, HR=hazard ratio, 95%CI=95% confidence interval

**Figure 3:**
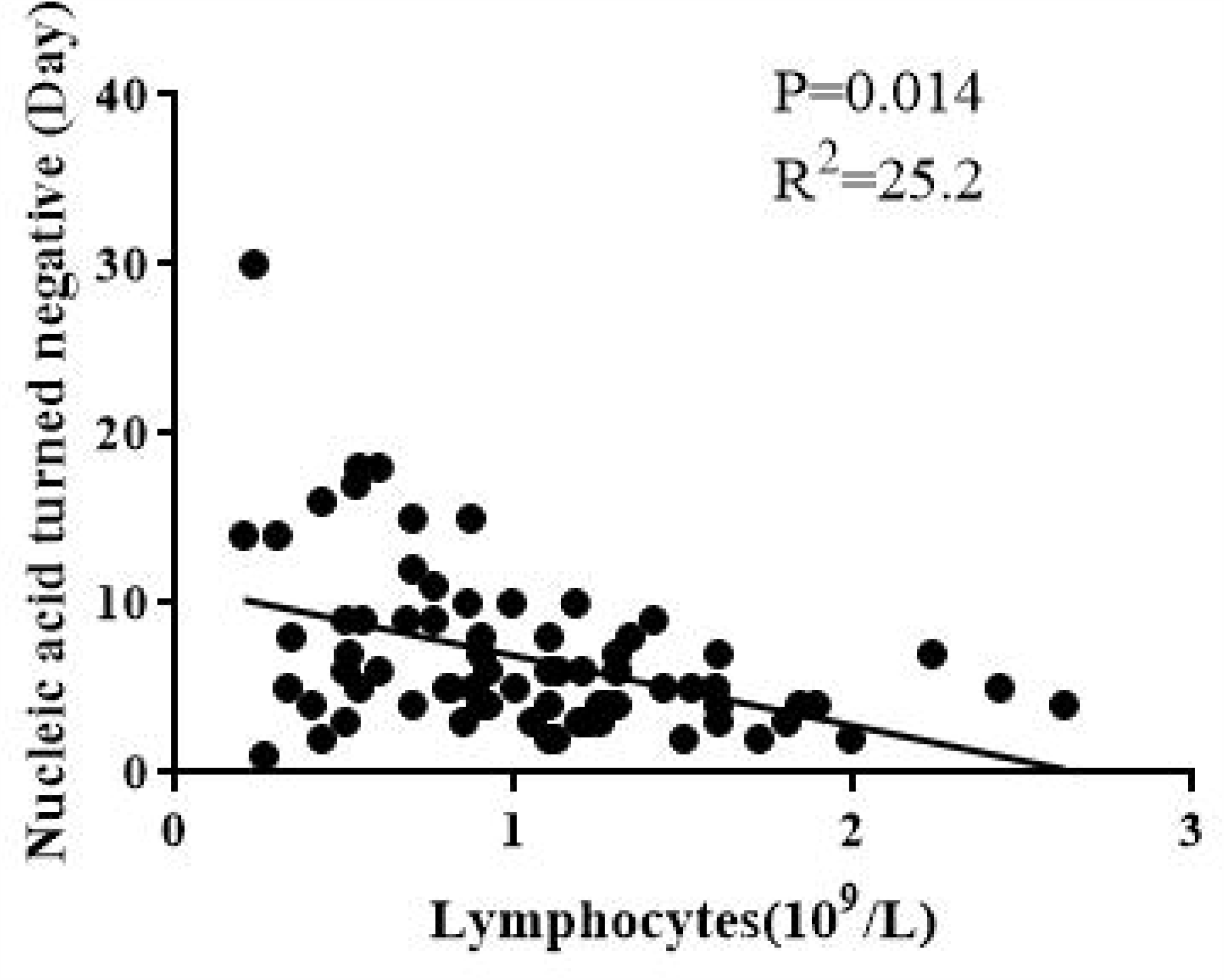
Correlations between: The days of nucleic acid test turned negative and lymphocytes;

**Figure 4:**
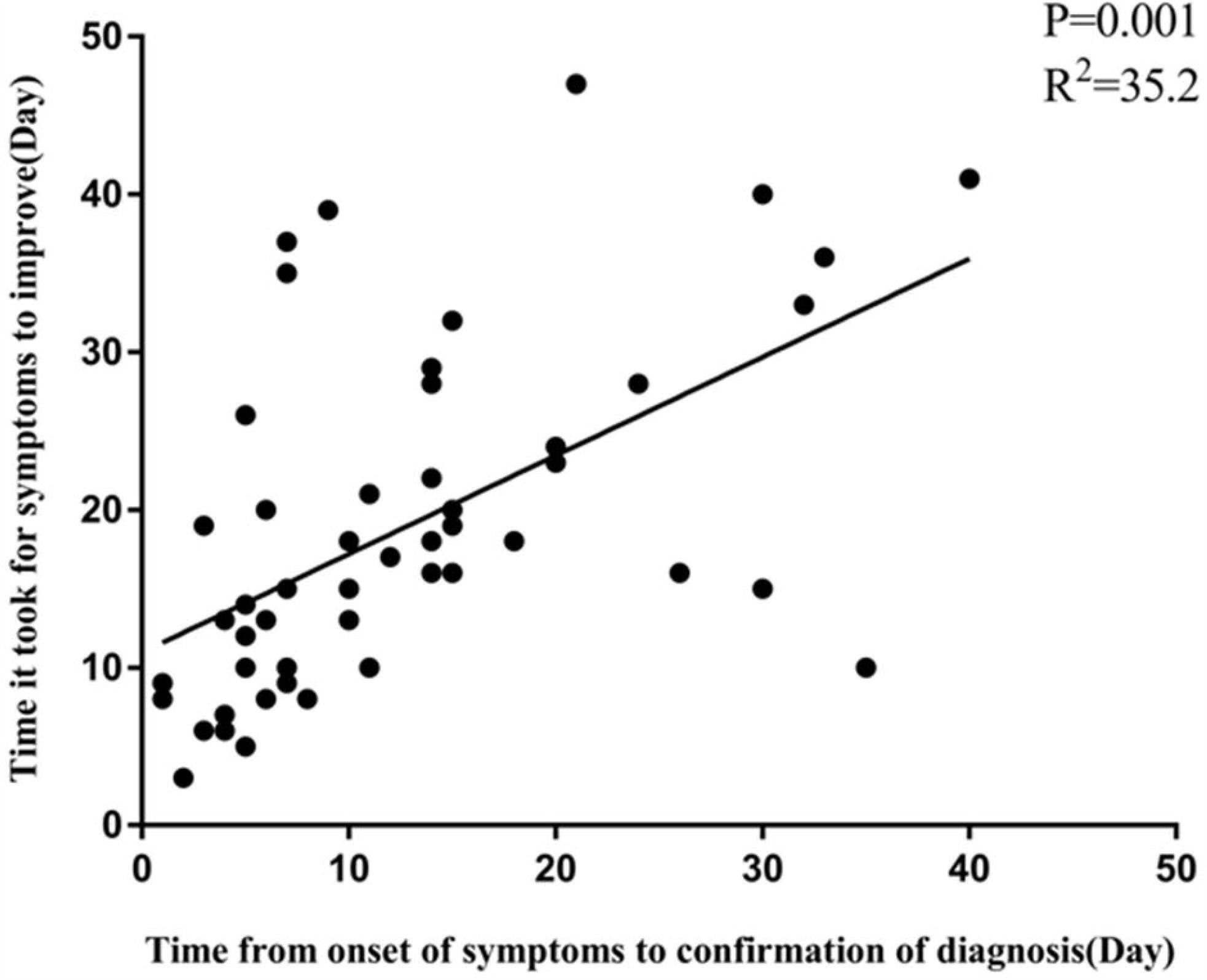
Correlations between: Time from onset of symptoms to confirmation of diagnosis and Time it took for symptoms to improve.

## 4. Discussion

This study summarizes the clinical characteristics, laboratory findings, dynamic changes in CT images, treatment, and prognosis of 169 COVID-19 patients from 5 hospitals in a western China city and in the city of disease onset, Wuhan. The study included mostly mild cases of COVID-19, which accounted for 88.2%. We analyzed the disease progression in three ways: number of days from positive COVID-19 nucleic acid test to negative test, number of days from onset of illness to improvement in symptoms, and improvement in second CT scan compared to the first. At present, no similar large sample data has been reported.

The study showed that the common symptoms of COVID-19 were fever, cough, sputum production, dyspnea, diarrhea, and myalgia. The main abnormalities in laboratory tests included that lymphocytopenia and increase in levels of CRP, LDH, and D-dimer, while PCT was normal. These findings are consistent with other literature reports.4,5,8,9 The most common comorbidities were hypertension, cardiovascular disease, and diabetes. Chronic obstructive pulmonary disease was rare. Both clinical symptoms and laboratory tests indicated that patients with comorbidities had worse course of the disease, with higher proportion of severe or critical disease and poorer prognosis. Patients with comorbidities manifested with more dyspnea, lower lymphocyte count, and higher LDH, compared with those who had no comorbidities. These were manifestations of COVID-19 disease, rather than worsening symptoms of the comorbidities due to infection with SARS-Cov-2. We also found that the proportion of critical type COVID-19 increased with age. Lower lymphocyte count and SpO2were found in a higher proportion of patients with severe or critical type. These findings suggest that healthcare providers should pay particular attention to age, comorbidities, lymphocyte count, and SpO2 in the diagnosis and treatment of COVID-19. These factors may predict the progression of the patient from moderate to severe or critical type.

Early CT findings showed that the lung lesions were mainly bilateral, mainly involved two or more lobes, and were mainly distributed in the peripheral zones of the lung. GGO, consolidation and mixed GGO and consolidation were the most common CT findings, while pleural effusion was rare. This is also consistent with previous literature reports.10 Through statistical analysis of lung CT imaging on different days in the course of the disease, we found the following progression: the early CT finding was GGO (1–7 days);the GGO lesion resolved or gradually evolved into consolidation shadow (1–2 weeks); consolidation resolved(2–3 weeks);then, linear opacity or reticular changes developed (4–6 weeks).The whole course took about a month. In the statistical analysis of the correlation between symptom improvement and pulmonary imaging changes in 36 patients, nearly half of the patients (16/36) showed resolution of GGO lesions and aggravation of consolidation lesions when the clinical symptoms improved. Histological Lung biopsy specimen showed interstitial mononuclear inflammatory infiltrates (dominated by lymphocytes), diffuse alveolar damage with cellular fibro-myxoid exudate, or hyaline membrane formation.11 This is different from what is seen in bacterial pneumonia. Therefore, the change in extent of GGO lesions can be used to predict the outcome of the disease, while consolidation shadow should not be used as aprognostic factor for the patients’ condition.

In this study, all patients had positive nucleic acid tests for pharyngeal swabs. Seven cases had positive nucleic acid test for stool samples, but only six cases had diarrhea. One patient tested positive for urine nucleic acid test, which is a rare report at present, and may suggest that SARS-Cov-2 may cause urinary tract infection.

Most of the patients in this study were treated with antiviral drugs, including lopinavir and tonavir. Currently, there are no research data that show that such drugs have a clear effect on COVID-19. In this study, we found that the lower the lymphocyte count, the more the number of days for nucleic acid test to change from positive to negative. The longer the duration from onset of illness to admission, the longer the duration before improvement of symptoms, and the shorter the duration from onset of illness to admission, the shorter the duration for CT image improvement. Comparing patients in Hubei and Guangxi, we found that patients from Hubei had more severe disease and more days from onset of illness to admission than those from Guangxi. All of these findings suggest that early isolation, early diagnosis, and early initiation of management can contribute to slowing down the progression and spread of COVID-19.

The present study has some limitations. First, due to the limited number of cases, some results need to be further validated with more patients. Second, the effect of antiviral agents and corticosteroidson COVID-19 needs further validation. Prospective studies should be performed to get more accurate results. Third, because this was a retrospective analysis, the time of CT imaging and nucleic acid test was not standardized and unified, t may have reduced the accuracy of the result. More cases need to be analyzed to obtain more information.

## 5. Conclusions

In this study, we found that older patients and patients with comorbiditieshad higher proportion of severe or critical type COVID-19. Lymphocyte count and and SpO2 at admission were associated with disease severity. Early isolation, early diagnosis, and early initiation of management can contribute to slowing down the progression and spread ofCOVID-19. These findings will be helpful for the diagnosis and treatment of COVID-19.

## Data Availability

All data in the manuscript is available

## Author Contributions

WZ made substantial contributions to the conception and design of the study; acquisition, analysis, and interpretation of the data; and drafting of the manuscript. XF, JH, CD and DQ made substantial contributions to the conception and design of the study; acquisition, analysis, and interpretation of the data; and critical revision of the manuscript for important intellectual content. XZ and JZ also gave final approval of the version to be published and agree to be accountable for all aspects of the work in ensuring that questions related to the accuracy and integrity of any part of the work are appropriately investigated and resolved. JZ gave final approval of the version to be published. WZ and XF participated in analysis and interpretation of the data and agrees to be accountable for all aspects of the work in ensuring that questions related to the accuracy or integrity of any part of the work are appropriately investigated and resolved. WZ and JZ conceived of the study, participated in its design, and helped to draft the manuscript. All authors read and approved the final manuscript.

## Acknowledgements

We would like to thank the three radiologists, Drs. Yanyan Shen, Peng Peng, Dong Deng, who helped with the analysis and interpretation of the imaging data.

## Ethical approval

This study was approved by the ethics committee associated with the Faculty of Medicine at The First Affiliated Hospital of Guangxi Medical University[2020(KY-E-06).]. Informed consent was done appropriately for subjects.

